# Unique proteome signatures in ICU patients with COVID-19 and delirium: an observational study

**DOI:** 10.1101/2024.12.09.24317145

**Authors:** Andreas Edel, Jayanth Sreekanth, Florian Kurth, Markus Ralser, Vadim Demichev, Michael Mülleder, Eric Blanc, Claudia Spies

## Abstract

**Background:** Delirium is common in COVID-19 intensive care unit (ICU) patients. Biomarkers for prediction, detection, and monitoring are missing. Unbiased omics analyses are warranted to gain a systems biology view on pathophysiology.

**Methods:** This prospective observational satellite study aims to investigate the proteome signatures of COVID-19 ICU patients, comparing those with delirium to those without. This study was conducted in ICUs of a university hospital between March 2020 and September 2021. ICU patients of legal age with a positive SARS-CoV-2 test were screened daily for oversedation and delirium. Blood samples were taken thrice a week. 457 samples were analyzed using data-independent acquisition mass spectrometry to determine protein levels. A mixed-effects regression model was developed to identify proteins significantly influenced by delirium, accounting for sex and age as confounders. This model also aimed to determine proteins that were either up- or downregulated in association with delirium. Additionally, an enrichment analysis was conducted to examine the biological pathways linked to these delirium-associated proteins.

**Results:** Out of 360 ICU patients, 69 were analyzed for protein profiling. Out of these 69 patients, 42 patients (60.9%) had delirium on ICU admission, and 27 (39.1%) did not. Based on the multivariate model, the analysis of 204 proteins unfolded 125 (61.3%) to be differentially expressed. In total, 80.8% (n=101) of these 125 proteins were associated with delirium. Of these, 10 proteins were uniquely associated with delirium and were not significant in the multivariate model (SERPING1, SERPINA7, HP, TGFBI, CD5L, IGHV3-7, IGHV1-46, IGHV3-15, IGHV3-23, and “IGHV4-34;IGHV4-38-2”). In the univariate model for delirium, six out of 111 significant proteins showed increased expression with a log2FC > 0.5: PIGR, MST1, LBP, CRP, SAA1, and “SAA1;SAA2”; while three showed decreased expression with a log2FC < - 0.5: HP, PPBP, and “HP;HPR”. The enrichment analysis of delirium-influenced proteins revealed three significant pathways: “Network map of SARS-CoV-2 signaling” (M42569/WP5115), “Acute inflammatory response” (M10617), and “Regulation of defense response” (M15277).

**Conclusion:** We identified a unique proteomic signature in COVID-19 ICU patients with delirium, including up- and downregulated proteins. These findings may provide potential biomarker candidates for the assessment of delirium risk and its underlying causes. These findings could be a further step towards the development of personalized, causative treatments for delirium and its monitoring in the ICU.

**Trial registration:** The study was retrospectively registered in the German Clinical Trials Register on May 13, 2020 (DRKS00021688).

## Background

Delirium is a condition of acute and fluctuating brain dysfunction characterized by disturbances of attention, awareness, and cognition and is not caused by any other neurocognitive diseases(1). Its pathophysiology and etiology are complex and multifactorial. The underlying etiologies of delirium are discussed with respect to metabolic, hypoxic, immune, and toxic encephalopathies (2). Hence, a model of predisposing and precipitating factors tries to explain its complexity(3). Viral infections, such as those caused by Severe Acute Respiratory Syndrome Coronavirus 2 (SARS-CoV-2), have been significant precipitating factors. SARS-CoV-2 triggers pro-inflammatory molecules in serum, causing delirium or encephalopathy as seen in other coronaviruses(4). The prevalence of delirium in an elderly patient cohort is around 34% (5), and it can increase to approximately 55% in ICU settings(6). Delirium is linked to worse outcomes, such as prolonged hospital treatment, readmission, and increased mortality(7). This has also been observed during the Coronavirus Disease 2019 (COVID-19) pandemic(8). Therefore, the World Health Organization (WHO) has urged clinicians to actively identify and manage delirium in hospitalized and critically ill patients(9). However, delirium remains underdiagnosed in hospitals(10). The diagnosis of delirium relies on cognitive tests such as the Confusion Assessment Method for the ICU (CAM-ICU)(11). Because of its limitations, researchers are increasingly searching for delirium biomarkers to predict, detect, and monitor delirium. Unbiased omics analyses are warranted to gain a systems biology view on pathophysiology without selecting markers based on prior knowledge. Here, high-throughput mass spectrometry offers a fast, cost-effective, and robust choice to profile proteomes at a scale(12). Previous studies investigating the proteomic signatures of patients with delirium mainly focused on a perioperative period(13). As a result, studies in the intensive care setting are lacking, even though delirium significantly impacts patient outcomes. To investigate biological processes in COVID-19 ICU patients with delirium, we focus on three primary objectives: 1) to identify significant proteins affected by delirium when sex and age are included as confounders; 2) to determine up- and downregulated proteins associated only with delirium; 3) to analyze these delirium-associated proteins using biological pathway analysis.

## Methods

### Study design and inclusion criteria

This is a satellite study of the prospective observational cohort study Pa-COVID-19 (ethics approval EA2/066/20) conducted at the Charité - Universitätsmedizin Berlin(14). This manuscript has been written following the guidelines of the STROBE statement for observational studies(15). All patients with a positive reverse transcriptase polymerase chain reaction (RT-PCR) to SARS-CoV-2 admitted to the university hospital between March 2020 and September 2021 and willing to give informed consent by themselves or their legal representatives were eligible for study inclusion. Clinical routine parameters and additional blood samples were collected into the study database. Out of this cohort, only patients older than 18 years and treated in the ICU between March 2020 and July 2020 were included in this satellite study. Patients who did not consent to blood sampling were excluded. The study was retrospectively registered (DRKS00021688), and its details have been previously reported(16).

### Variables

Baseline characteristics of age, sex, Body Mass Index (BMI), comorbidities, and ventilation status were obtained from the patient’s medical records. The Sequential Organ Failure Assessment (SOFA) score was selected without the Glasgow Coma Scale (GCS) domain. Sex and age were considered potential delirium confounders in further statistical analysis(17–19). For the pain assessment, the Numerical Rating Scale (NRS), Critical Care Pain Observation Tool (CPOT), and Behavioral Pain Scale (BPS) were collected and documented at least every eight hours according to local standard operating procedure (SOP).

As the primary outcome parameter, protein expression profiling from serum samples was performed using an untargeted mass spectrometry-based method as previously described(20). Blood samples were collected three times a week during hospitalization (up to 14 days) and, when feasible, at intervals of 6 weeks, 3 months, 6 months, and 12 months. The plasma proteomics data obtained through data-independent acquisition and label-free quantitation was processed as previously described(20). We retained the UniProt gene names for proteins, and the processed data included protein groups. The protein groups did not necessarily include paralogues, but sometimes just unrelated proteins sharing a peptide. The term “protein” includes individual proteins and protein groups.

### Delirium screening

In this analysis, delirium was diagnosed during the ICU treatment by a positive delirium screening using the Confusion Assessment Method for the ICU (CAM-ICU). Delirium includes oversedation, which was defined as a Richmond Agitation Sedation Scale (RASS) < −2. In accordance with the local SOP, this screening was performed and documented in patients’ charts at least every eight hours. After discharge from the ICU, the follow-up examination reports were manually reviewed for delirium.

### Bias

Unconscious patients were initially enrolled through an approved deferred consent process to minimize the potential selection bias. To identify delirium at the first presentation on admission to the ICU, we considered the first initial delirium screening. Given the fluctuating nature of delirium and the daily summarization of delirium status, potential biases may arise in exploring connections between the proteome and delirium, because we defined delirium to be positive if at least one positive screening occurred during the treatment day. Also, the imputed protein expression may not reflect the actual distribution.

### Statistical methods

Descriptive statistics were performed on patient characteristics obtained from the electronic health records, and the summary stratified according to delirium status at ICU admission are presented as counts with percentage (%) and median with interquartile range (IQR) for categorical and continuous variables, respectively. Continuous variables, except age, are rounded to one decimal place. All protein measurements were log2-transformed, and proteins with >20% missing values were discarded. As further analysis cannot handle missing values, the remaining proteins (with <20% missing values) were imputed using the minimum value of each protein. The expression variation of proteins influenced by each variable of interest was assessed using the *dream* function in *variancePartition* (v1.24.1) package in R(21, 22).

### Mixed-effects model including multiple covariates

To identify significantly up- and downregulated proteins, a mixed-effects regression model with individual patients as a random effect and delirium as a fixed effect was created. Age and sex were included as fixed effects to account for potential confounders. The resulting model was as follows: *∼ delirium + age + sex + (1 | patient ID)*. Delirium was considered a fixed effect, neglecting the variation in delirium status within individuals. The correlation between delirium, age, and sex was reviewed using canonical correlation analysis (CCA) embedded in the *variancePartition* (v1.24.1) package. Additional analyses were performed to determine all significantly up- and downregulated proteins for each covariate by defining the coefficients separately. P-values obtained using eBayes method(23) were corrected for multiple testing using the Benjamini-Hochberg false discovery procedure(24). We only deemed proteins with adjusted p-values below 0.05 to have significant differential expression.

### Enrichment analysis on proteins differentially expressed according to delirium

For further enrichment analysis, we only focused on proteins obtained when delirium was the only model coefficient. Screening of proteins based on their fold-change was not performed. We distinguished the proteins having log2FC ≥ 0 as upregulated and the ones with log2FC < 0 as downregulated. Without restricting the enrichment analysis to proteins with adjusted p-values below 0.05, we considered the complete list of proteins (this also includes protein groups) ranked according to B-statistic (default ranking) for the enrichment analysis. Two custom gene sets of Homo sapiens: GO Biological Process and WikiPathways were retrieved using *msigdbr* (v7.5.1) R package(25). We identified the potential pathways using a second-generation algorithm called the CERNO test included in the *tmod* (v0.50.13) package(26). Only the gene sets (pathways) having greater than 15 hits were retained, as this ensured the filtering of no and very few hits. After that, the p-values were re-computed for multiple testing using the Benjamini-Hochberg method. The gene sets with adjusted p-values below 0.05 were considered statistically significant.

The data analyses were performed, and images were generated in R (v4.1.2) using *tidyverse* (v1.3.2)(27), *dplyr* (v1.0.10)(28), *ggplot2* (v3.4.0)(29), *ggVennDiagram* (v1.2.3)(30), *EnhancedVolcano* (v1.12.0)(31), and *ComplexHeatmap* (v2.10.0)(32) packages.

## Results

### Descriptive statistics

Between March 2020 and September 2021, 1,699 patients with SARS-CoV-2 infection were treated in the ICUs of the Charité – Universitätsmedizin Berlin. Among these, 360 patients were included in the Pa-COVID-19 study. Of these, 69 patients had blood samples collected for proteomics analysis within the defined time period and were consequently included in this satellite study (Figure 1). 42 patients (60.9%) had delirium on their ICU admission, and 27 (39.1%) did not. The majority of patients with delirium on ICU admission were oversedated (n=33, 78.6%; 6) vs. 8 (7–12)]. The descriptive analysis of the cohort is summarized in Table 1. An explanatory map of sample collection and delirium status is presented as a heatmap in Supplementary Figure 1.

**Figure 1:**
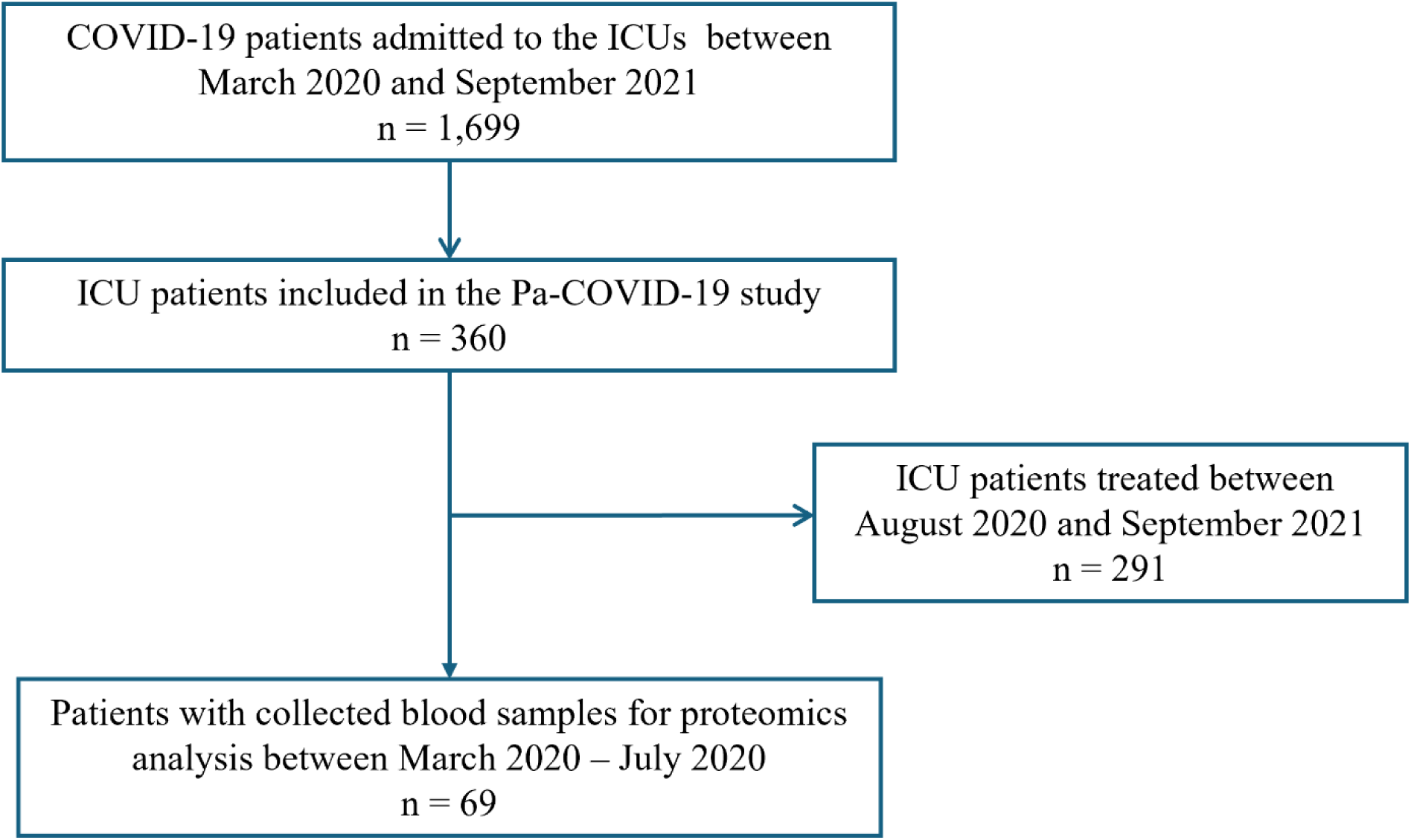
Flow chart of patient selection process in this study.

**Figure 2:**
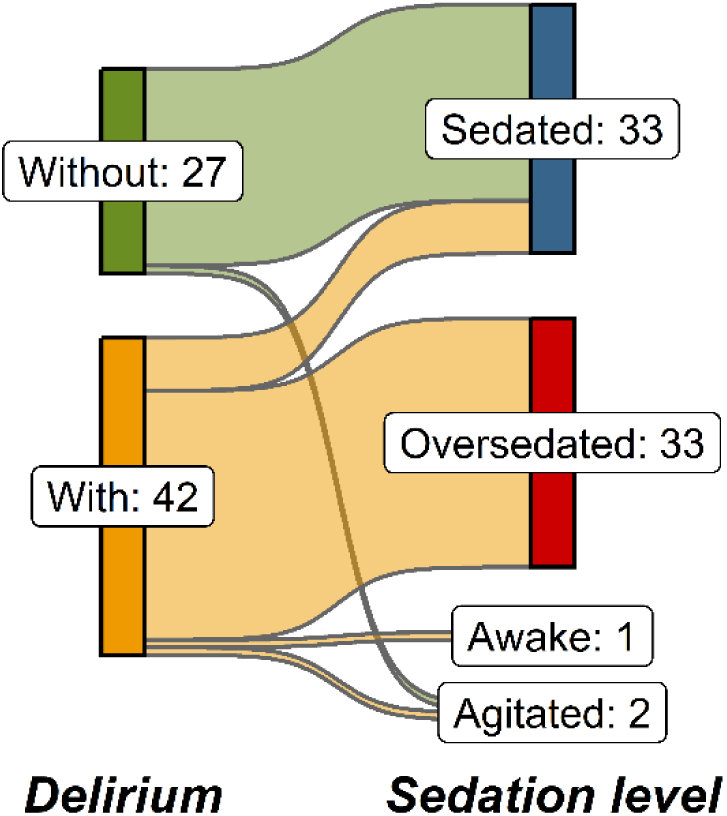
Sedation level in patients with and without delirium. Sankey plot displaying the sedation level in patients with and without delirium on ICU admission. Sedation level is defined on the Richmond Agitation-Sedation Scale (RASS); Agitated: RASS > 0, Awake: RASS = 0, Sedated: RASS −1 and −2, Oversedated: RASS < −2.

**Table 1:**
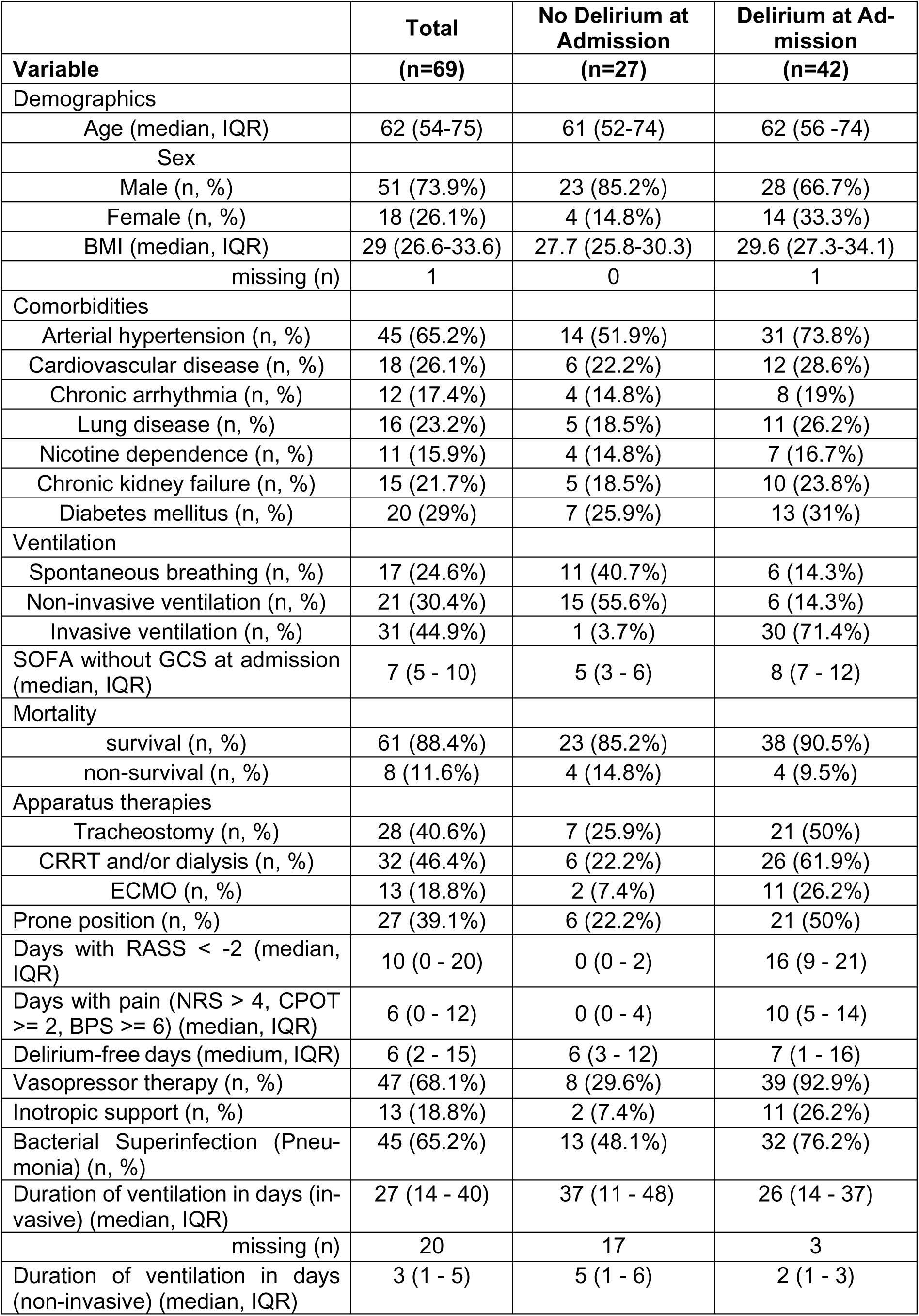
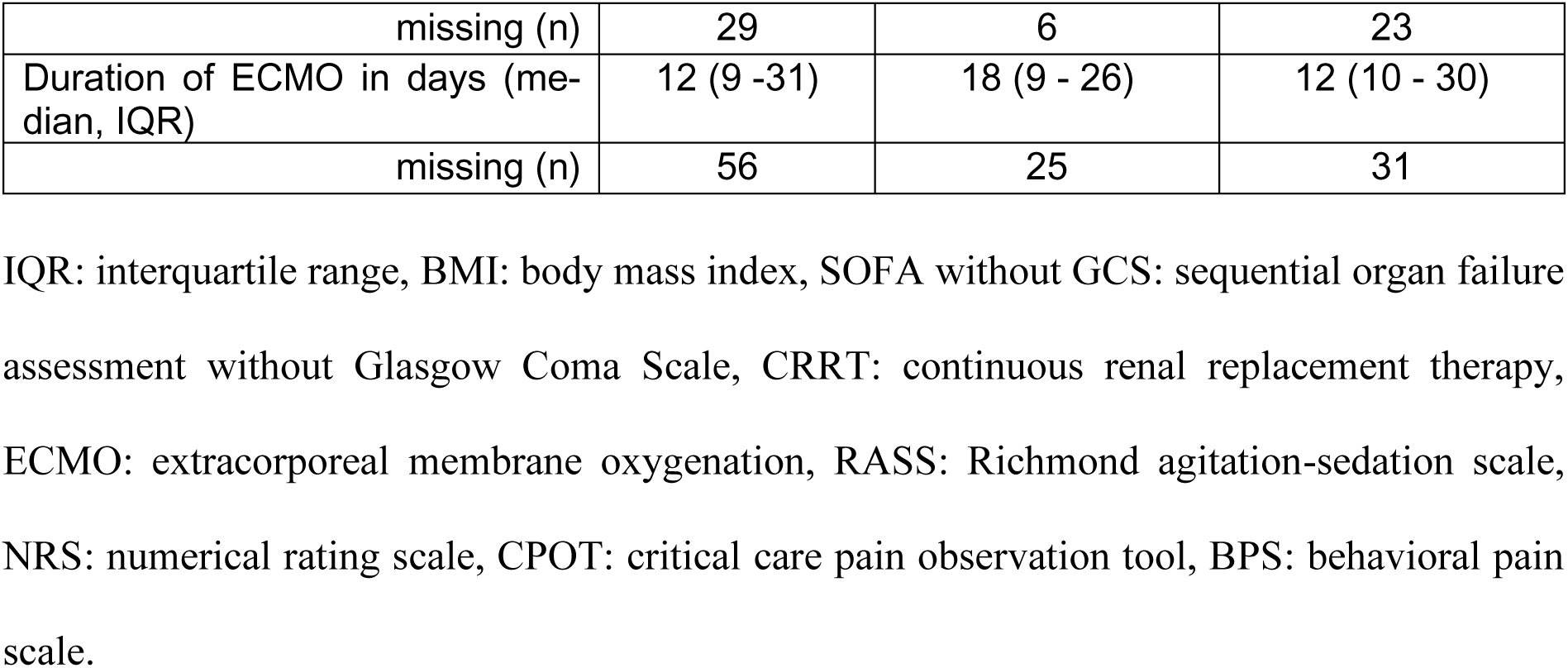
Patient information of all included subjects at their initial ICU admission.

|  | Total | No Delirium at Admission | Delirium at Admission |
| --- | --- | --- | --- |
| Variable | (n=69) | (n=27) | (n=42) |
| Demographics |  |  |  |
| Age (median, IQR) | 62 (54-75) | 61 (52-74) | 62 (56 -74) |
| Sex |  |  |  |
| Male (n, %) | 51 (73.9%) | 23 (85.2%) | 28 (66.7%) |
| Female (n, %) | 18 (26.1%) | 4 (14.8%) | 14 (33.3%) |
| BMI (median, IQR) | 29 (26.6-33.6) | 27.7 (25.8-30.3) | 29.6 (27.3-34.1) |
| missing (n) | 1 | 0 | 1 |
| Comorbidities |  |  |  |
| Arterial hypertension (n, %) | 45 (65.2%) | 14 (51.9%) | 31 (73.8%) |
| Cardiovascular disease (n, %) | 18 (26.1%) | 6 (22.2%) | 12 (28.6%) |
| Chronic arrhythmia (n, %) | 12 (17.4%) | 4 (14.8%) | 8 (19%) |
| Lung disease (n, %) | 16 (23.2%) | 5 (18.5%) | 11 (26.2%) |
| Nicotine dependence (n, %) | 11 (15.9%) | 4 (14.8%) | 7 (16.7%) |
| Chronic kidney failure (n, %) | 15 (21.7%) | 5 (18.5%) | 10 (23.8%) |
| Diabetes mellitus (n, %) | 20 (29%) | 7 (25.9%) | 13 (31%) |
| Ventilation |  |  |  |
| Spontaneous breathing (n, %) | 17 (24.6%) | 11 (40.7%) | 6 (14.3%) |
| Non-invasive ventilation (n, %) | 21 (30.4%) | 15 (55.6%) | 6 (14.3%) |
| Invasive ventilation (n, %) | 31 (44.9%) | 1 (3.7%) | 30 (71.4%) |
| SOFA without GCS at admission (median, IQR) | 7 (5 - 10) | 5 (3 - 6) | 8 (7 - 12) |
| Mortality |  |  |  |
| survival (n, %) | 61 (88.4%) | 23 (85.2%) | 38 (90.5%) |
| non-survival (n, %) | 8 (11.6%) | 4 (14.8%) | 4 (9.5%) |
| Apparatus therapies |  |  |  |
| Tracheostomy (n, %) | 28 (40.6%) | 7 (25.9%) | 21 (50%) |
| CRRT and/or dialysis (n, %) | 32 (46.4%) | 6 (22.2%) | 26 (61.9%) |
| ECMO (n, %) | 13 (18.8%) | 2 (7.4%) | 11 (26.2%) |
| Prone position (n, %) | 27 (39.1%) | 6 (22.2%) | 21 (50%) |
| Days with RASS < -2 (median, IQR) | 10 (0 - 20) | 0 (0 - 2) | 16 (9 - 21) |
| Days with pain (NRS > 4, CPOT >= 2, BPS >= 6) (median, IQR) | 6 (0 - 12) | 0 (0 - 4) | 10 (5 - 14) |
| Delirium-free days (medium, IQR) | 6 (2 - 15) | 6 (3 - 12) | 7 (1 - 16) |
| Vasopressor therapy (n, %) | 47 (68.1%) | 8 (29.6%) | 39 (92.9%) |
| Inotropic support (n, %) | 13 (18.8%) | 2 (7.4%) | 11 (26.2%) |
| Bacterial Superinfection (Pneumonia) (n, %) | 45 (65.2%) | 13 (48.1%) | 32 (76.2%) |
| Duration of ventilation in days (invasive) (median, IQR) | 27 (14 - 40) | 37 (11 - 48) | 26 (14 - 37) |
| missing (n) | 20 | 17 | 3 |
| Duration of ventilation in days (non-invasive) (median, IQR) | 3 (1 - 5) | 5 (1 - 6) | 2 (1 - 3) |
| missing (n) | 29 | 6 | 23 |
| Duration of ECMO in days (median, IQR) | 12 (9 -31) | 18 (9 - 26) | 12 (10 - 30) |
| missing (n) | 56 | 25 | 31 |

IQR: interquartile range, BMI: body mass index, SOFA without GCS: sequential organ failure assessment without Glasgow Coma Scale, CRRT: continuous renal replacement therapy, ECMO: extracorporeal membrane oxygenation, RASS: Richmond agitation-sedation scale, NRS: numerical rating scale, CPOT: critical care pain observation tool, BPS: behavioral pain scale.

### Observation of protein expression in the multivariate mixed model

Exploration of missing values revealed that 127 (38.4%) out of 331 proteins had more than 20% missing values. The associated difference in the plasma protein concentration (including protein groups) is reported by having 457 samples and 204 proteins (<20% missing values). Before calculating the regression model, the canonical correlation analysis revealed a weak correlation with age and sex, whereas delirium did not correlate with age and sex.

The results of the multivariate regression model with delirium, age, and sex as covariates revealed 125 (61.3%) significantly differentially expressed proteins (adjusted p-value < 0.05). Of these, 10 proteins were differently expressed in the multivariate model, but not when the coefficients were defined individually (Supplementary File 1). Individual analysis for each covariate showed 32 proteins significant for age and only one for sex. Most proteins (n = 101; 80.8%) were affected by delirium. The Pregnancy Zone Protein (PZP) (log2FC: −0.86 for males), identified as significant in relation to sex, was also found to be significant for delirium (Figure 3; Supplementary File 2).

**Figure 3:**
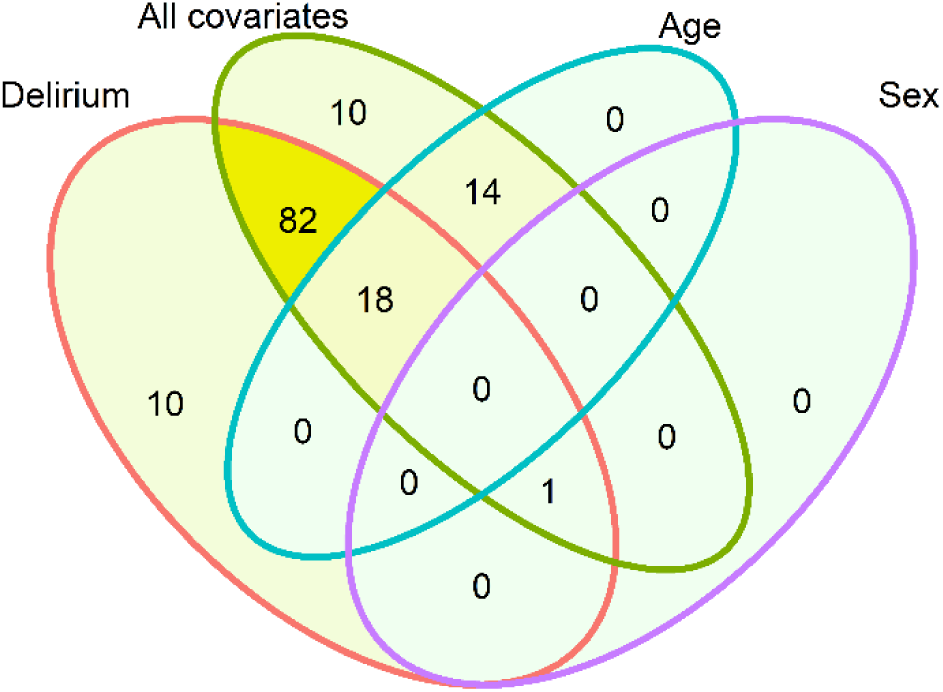
Analysis of differentially expressed proteins affected by delirium, age and sex. Differentially expressed protein numbers are shown using a Venn diagram to distinguish the significant proteins affected by delirium, age, sex, and altogether.

Of the 32 proteins significant for age, 18 were common with respect to delirium and the remaining 14 were distinct to age (Supplementary File 1). The proteins significant for age and delirium are given in Table 2, including their direction of expression. We observe that the following proteins exhibit opposite directions of change: A2M, FETUB, KNG1, PRG4, HABP2, “A2M; PZP”.

**Table 2:** Comparison of expression directions of common significant proteins (n=18) between delirium and age.

| **Proteins**<br>**(UniProt gene names)** | **Delirium (Positive)** | **Age** |
| --- | --- | --- |
| A2M | Downregulated | Upregulated |
| A2M; PZP | Downregulated | Upregulated |
| AHSG | Downregulated | Downregulated |
| APOM | Downregulated | Downregulated |
| C7 | Upregulated | Upregulated |
| CD14 | Upregulated | Upregulated |
| CST3 | Upregulated | Upregulated |
| F2 | Downregulated | Downregulated |
| F12 | Downregulated | Downregulated |
| FETUB | Upregulated | Downregulated |
| HABP2 | Upregulated | Downregulated |
| IGFALS | Downregulated | Downregulated |
| IGHA1 | Upregulated | Upregulated |
| ITIH3 | Upregulated | Upregulated |
| ITIH4 | Upregulated | Upregulated |
| KLKB1 | Downregulated | Downregulated |
| KNG1 | Upregulated | Downregulated |
| PRG4 | Upregulated | Downregulated |

A2M: Alpha-2-Macroglobulin, A2M; PZP: Alpha-2-Macroglobulin and Pregnancy Zone Protein (PZP), AHSG: Alpha-2-HS-Glycoprotein (Fetuin-A), APOM: Apolipoprotein M, C7: Complement Component 7, CD14: Cluster of Differentiation 14, CST3: Cystatin C, F2: Coagulation Factor II (Prothrombin), F12: Coagulation Factor XII (Hageman Factor), FETUB: Fetuin-B, HABP2: Hyaluronan-Binding Protein 2, IGFALS: Insulin-Like Growth Factor-Binding Protein, Acid Labile Subunit, IGHA1: Immunoglobulin Heavy Constant Alpha 1, ITIH3: Inter-Alpha-Trypsin Inhibitor Heavy Chain 3, ITIH4: Inter-Alpha-Trypsin Inhibitor Heavy Chain 4, KLKB1: Kallikrein B, Plasma (Fletcher Factor) 1, KNG1: Kininogen-1, PRG4: Proteoglycan 4.

### Differentiation of up- and downregulated proteins in patients with and without delirium

In the univariate model with delirium as the coefficient, 111 out of the 204 proteins (54.4%) were significantly different, with a median log2FC of 0.108 (Figure 4 and Supplementary File 2). Of these, 10 proteins (9%) were uniquely associated with delirium and were not significant in the multivariate model. These proteins were SERPING1, SERPINA7, HP, TGFBI, CD5L, IGHV3-7, IGHV1-46, IGHV3-15, IGHV3-23, and “IGHV4-34;IGHV4-38-2” (Supplementary File 1). Among the 111 significant proteins, 66 (59.5%) were found to be increased (log2FC ≥ 0), and 45 (40.5%) were seen to be decreased (log2FC < 0). Six proteins (4.5%) had an increased expression with a log2FC > 0.5: PIGR, MST1, LBP, CRP, SAA1 and “SAA1;SAA2”. In contrast, three proteins (2.7%) showed a decreased expression with a log2FC < −0.5: HP, PPBP, and “HP;HPR” (Figure 4; Supplementary File 2).

**Figure 4:**
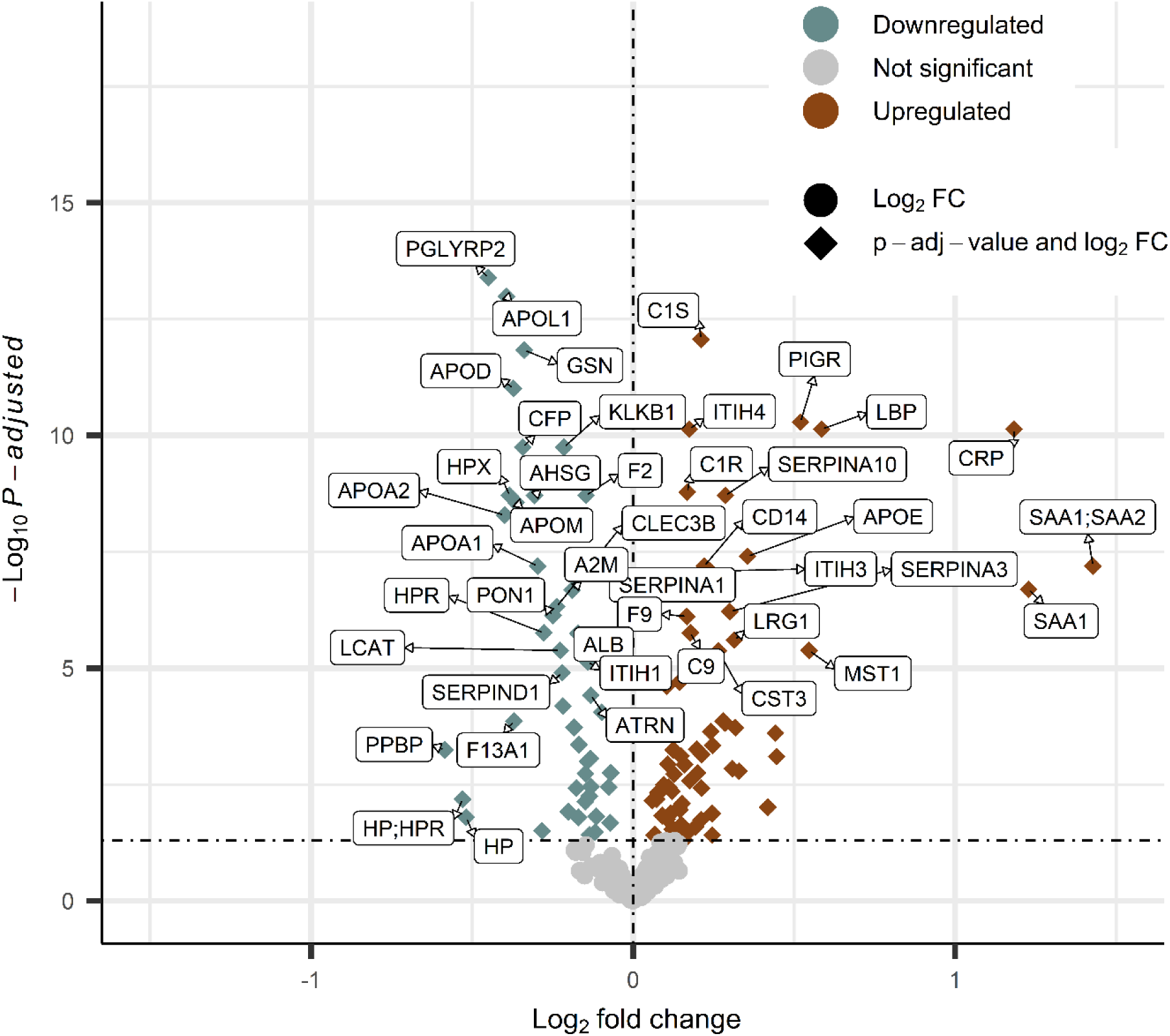
Visualization of the protein expression. Volcano plot when delirium was defined as the only model coefficient. Upregulated proteins are colored brown, and downregulated proteins are colored turquoise.

### Gene set enrichment analysis of delirium-influenced proteins

The gene set enrichment analysis revealed the following three significant pathways (Figure 5): “Network map of SARS-CoV-2 signaling” (M42569/WP5115), “Acute inflammatory response” (M10617), and “Regulation of defense response” (M15277). The majority of proteins included in the pathway “Regulation of defenses response” are incorporated in the pathway “Acute inflammatory response” except for the two proteins SERPING1, and HPX. Proteins with their corresponding pathways are reported in Supplementary File 3.

**Figure 5:**
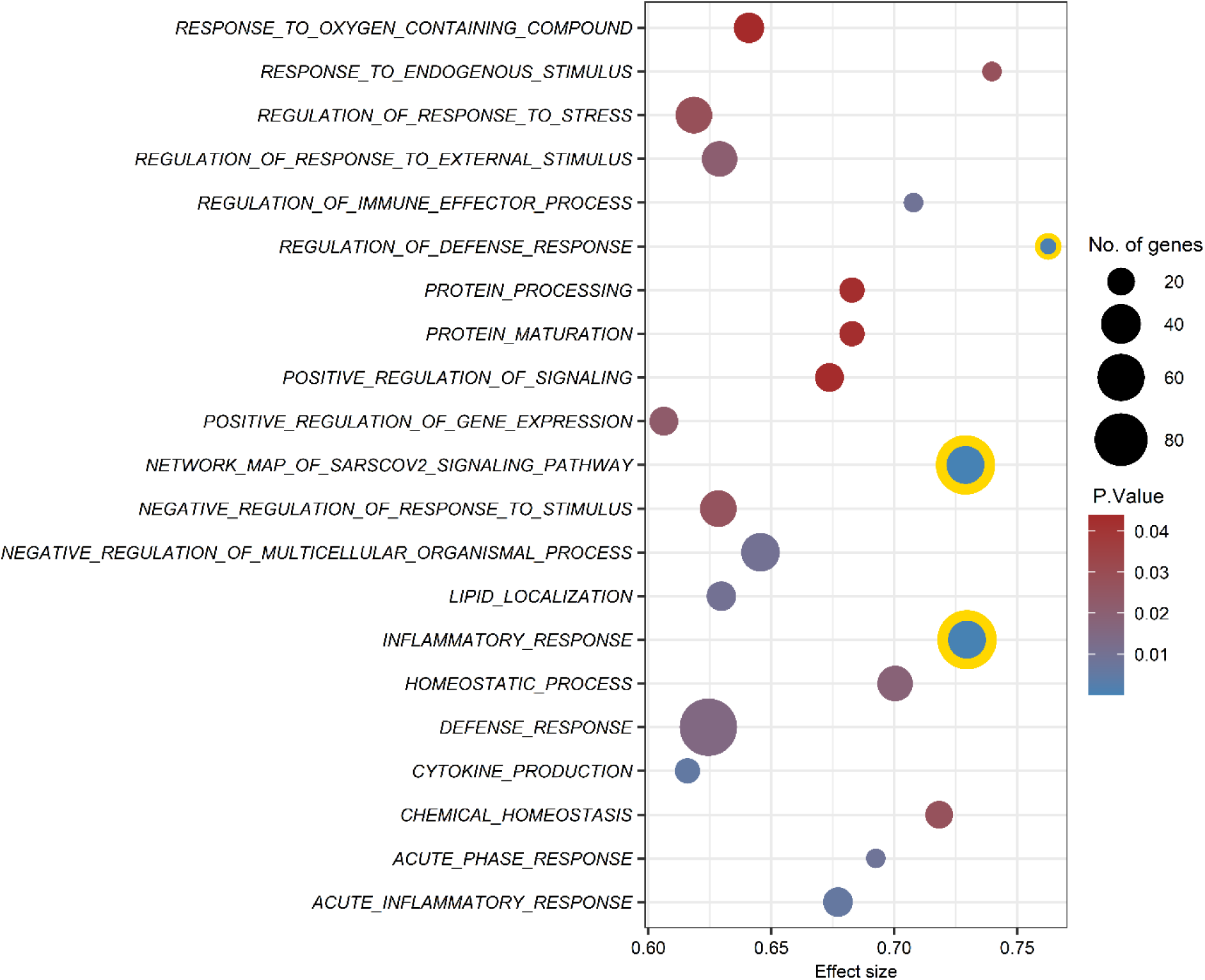
Visualization of enriched gene set associated with delirium. Portrayal of the potential gene sets found to be enriched for delirium. The plots’ bubbles are sized according to the number of genes and colored according to the p-values. The gene sets with re-adjusted p-values less than 0.05 have been highlighted in yellow.

## Discussion

### Protein expression differences in the multivariate model

We conducted this study to investigate whether COVID-19 ICU patients with delirium would have different protein expression levels compared to patients without delirium. For this, we selected consecutive blood samples of patients and analyzed them using mass spectrometry. Our multivariate regression model identified the majority of the proteins (n = 101; 80.8%) as being affected by delirium, although age and sex may also have some effect. This supports the potential hypothesis that the delirium status may affect the proteome. However, further research is needed on the ICU patient cohort. Currently, only a few perioperative studies involving proteomic analyses of different human body fluids(33–38) are available, which show similar results. Here, Oren et al. discovered a difference in proteome signature between differing age groups in postoperative patients with delirium(19). We also identified proteins that are influenced by age and partially overlap with those regulated by delirium. This may support the theory that an age-related mechanism is also involved in pathogenesis of delirium(19). The proteomic analysis of mass spectrometry is typically performed on ICU patients’ samples for specific diseases such as COVID-19(39, 40), sepsis(41), or neuro-trauma(42, 43). By combining the focus on acute viral infection and delirium, our study group was the first to investigate the proteome in this particular cohort.

### Proteins shared by both delirium and age

While comparing the multi- and univariate mixed-effect models, we identified six proteins - A2M, FETUB, KNG1, PRG4, HABP2, “A2M;PZP” - whose direction of expression differed between delirium and age. Alpha-2-Macroglobulin (A2M) plays a crucial role in regulating the immune response during infection. In our analysis, A2M and “A2M;PZP” were downregulated in patients with delirium and upregulated in older patients. As summarized in the review by Vandooren et al., A2M exhibits direct antibacterial properties through its protease activity and enhances the immune cell functions, including phagocytosis and antimicrobial capacity(44). Consequently, downregulated A2M levels may indicate an excessive infection status, which is frequently associated with a delirious state. As a protease inhibitor and cytokine inhibitor, A2M is also involved in the pathophysiology of Alzheimer’s disease by regulating the clearance of beta-amyloid deposits(45). Higher A2M concentrations have been linked to increased levels of neuronal injury markers, suggesting a potential association with delirium(46). Tunstall et al. previously described an age-dependent increase of A2M levels in healthy men (47), which may also be reflected in our results. Pregnancy zone protein, which is present in late pregnancy serum, has a similar structure and function to that of A2M(48).

Fetuin B (FETUB) is a member of the fetuin family, closely related to fetuin A, and is primarily secreted by the liver. In our study, FETUB showed an upregulated expression in patients with delirium and a downregulated expression in older patients. FETUB is involved in various metabolic processes, including osteogenesis, bone resorption, and insulin resistance in response to systemic inflammation(49, 50). To the best of our knowledge, our study is the first to report a possible association between fetuin B and delirium. Although its role in infections or sepsis is not well understood, animal studies suggest that it may play an additional role in these conditions(51).

In our analysis, Kininogen 1 (KNG1) was upregulated in patients with delirium and downregulated in older patients. KNG1 serves as a precursor protein for low molecular weight kininogen (LMWK) and high molecular weight kininogen (HMWK), with HMWK playing a role in blood coagulation and the kallikrein-kinin-system (KKS)(52). Given that the KKS pathway is significantly affected in COVID-19(53), this could explain the upregulation of KNG1 in seriously ill patients with delirium. Additionally, kininogen levels vary with age, being twice as high in adults compared to newborns(54). In a small study, researchers demonstrated that kininogen levels decrease with aging, which may also explain our observed downregulation of KNG1 expression in older patients.

In our study, Proteoglycan 4 (PRG4) exhibited opposite expression patterns: it was upregulated in patients with delirium and downregulated in older patients. PRG4, also known as lubricin, was initially identified in synovial fluid for its lubricating properties(55). Additionally, lubricin exhibits anti-inflammatory characteristics, reducing cytokine production by acting as an antagonist on Toll-like receptors, as summarized in the review by Richendrfer and Jay(56). Contrary to our findings of upregulated expression in patients with delirium and potentially seriously ill patients, previous studies have shown that lubricin expression is downregulated by pro-inflammatory cytokines such as interleukin-1 and tumor necrosis factor-alpha (57).

Finally, we identified opposite expression pattern for hyaluronan binding protein 2 (HABP2) in relation to delirium and age. Circulating hyaluronic acid (HA) in blood has been associated with lung injury and SOFA score in patients with ARDS(58). Consequently, it is assumed that HA plays a role in the lung pathophysiology of COVID-19(59). HABP2, also known as factor VII activating protease (FSAP), was initially identified for its ability to bind HA. This protein is also connected to the coagulation and fibrinolysis systems(60). Moreover, HABP2 is thought to negatively regulate vascular integrity in mice and in vitro models (61). Given its involvement in the inflammatory process, HABP2 may also reflect an increased inflammatory status in COVID-19 patients with delirium.

### Differentiation of up- and downregulated proteins in patients with delirium

In our study, we identified 10 proteins that were uniquely influenced by delirium and not significantly present in the multivariate model. These proteins include SERPING1, SERPINA7, HP, TGFBI, CD5L, IGHV1-46, IGHV3-7, IGHV3-15, IGHV3-23, and “IGHV4-34;IGHV4- 38-2”.

Plasma protease C1 inhibitor (C1INH), encoded by Serpin Family G Member 1 (SERPING1) gene, controls, via inhibitory inflammatory pathways, the permeability of the blood-brain barrier, and this can lead to cognitive impairments. Farfara et al. showed this pathophysiological pathway in a knock-out mice model(62). In contrast, Wiredu et al. could not find significant difference for C1INH in patients with delirium undergoing cardiac surgery(63). Contrary results were shown by Poljak et al. They investigated the proteomic profile of cerebrospinal fluid in 32 geriatric patients, having delirium, Alzheimer’s disease, or no cognitive impairment. The measured with the Mini-Mental State Examination (MMSE) score(64). In COVID-19, the level of C1INH is altered. Thus, Medjeral-Thomas et al. found that the level was significantly decreased in dialysis patients with severe COVID-19 compared to patients with dialysis without COVID-19 [difference of means 168.6; 95% CI (72.84 – 264.4); p = 0.003]. This decrease of C1INH in COVID-19 patients is assumed to be caused by strong connectivity between viral interactomes and C1INH(65).

Thyroxin-binding globulin (TBG), which is encoded by the Serpin Family A Member 7 (SERPINA7) gene, plays a major role in transporting the thyroid hormones thyroxine (T4) and triiodothyronine (T3) in plasma. In critically ill patients, thyroid function is altered. Here, T3 and T4 are reduced, and thyroid-stimulating hormone (TSH) is normal or slightly decreased. This is known as non-thyroidal illness (NTI)(66). Similar changes could be seen in the study of Chen et al. and Clausen et al., where patients with COVID-19 had a significantly lower concentration of thyroid hormones and these lower levels were also associated with mortality(67, 68). NTI is also hypothesized to be linked with postoperative delirium, as these thyroidal alterations were also observed in patients receiving major surgery(69). As TBG has a high affinity to T3 and T4, it has been suggested that the affinity of TBG to thyroid hormones is reduced in NTI. This reduced binding capacity is associated with a decreased concentration of total T4 but an increased concentration of free T4(70). However, in critically ill patients, this relationship is controversial, critical illness is often associated with decreased levels of free T4(71). Higher concentrations of free T4 have been linked with dementia(72). Nonetheless, a direct link between TBG and delirium has not been established.

As an acute-phase protein, produced by the liver, haptoglobin (HP) has been linked to cognitive function, thus HP (73) and HPR(74) have antimicrobial properties and can bind free hemoglobin after hemolysis(75). In contrast to our detected decreased level of HP, Westoff et al. found in their study on patients with postoperative delirium that the level of HP was increased in the cerebrospinal fluid (76). Additionally, an increased concentration of HP was found in the cognitive-related brain regions in rats(77) and was found to be significantly higher in patients with mild cognitive dysfunction and dementia(78). Moreover, patients in the ICU suffering from severe COVID-19 may develop delirium along with respiratory insufficiency. In some cases, extracorporeal membrane oxygenation (ECMO) therapy is required to support the patient’s respiratory function. However, it is essential to note that ECMO-induced hemolysis may lead to decreased levels of HP and HRP, as these proteins are depleted in this patient cohort.

The production of transforming growth factor-beta-induced protein (TGFBI) is stimulated by transforming growth factor (TGF)-β and secreted by several cell types, including macro- and microglia. TGF-β is predominantly involved in wound healing processes and plays a crucial role in the development of glial scar following brain injury (79). Since TGFBI is also secreted by other cells during the wound healing process(80), it is challenging to pinpoint the specific source in patients with severe COVID-19.

The main macrophage-secreted CD5 antigen-like (CD5L) protein plays multiple roles in inflammatory processes, including those related to infections, sterile inflammation, and autoimmune conditions. Specifically, CD5L acts as a regulatory protein in sepsis, ARDS, and bacterial infections(81). These conditions can also influence the development of delirium, complicating the identification of the underlying causes.

IGHV1-46, IGHV3-7, IGHV3-15, IGHV3-23, and “IGHV4-34;IGHV4-38-2” represent different variable regions of the heavy chain of immunoglobin (Immunoglobulin Heavy Chain variable region, IGHV). Among other IGHVs, D’alessandro et al. identified IGVH-3 as an increased protein in SARS-CoV-2 positive patients compared to controls(82). In patients with COVID-19, the diversity of the IGHV region is reduced. Gao et al. found that IGHV1-18 and IGHV1-69 were upregulated, while IGHV3-15, IGHV3-72, IGHV4-28, and IGHV7-81, which have lower frequencies (<2%), were significantly downregulated in COVID-19 patients. Conversely, IGHV4/OR15-8, IGHV4/OR15-7, IGHV3/OR16-15, IGHV3-54, and IGHV3-16 were significantly upregulated(83). There is limited evidence regarding the role of immunoglobins in SARS-CoV-2-induced neurological impairments. Muccioli et al. reported that five COVID-19 patients with encephalopathy, who received intravenous immunoglobin therapy, completely recovered from their neuropsychiatric symptoms(84). The Fc-portion, as part of the IGHV, has immune-regulating effects on pro-inflammatory cytokines(85) and may mediate the efflux of IgG through the blood-brain barrier, as shown in animal experiments(86). These preliminary investigations can only generate hypotheses regarding the influence of IGVH on brain function and its disruption in the form of delirium. However, further validation is required to determine whether these 10 proteins are directly impacted by delirium, as their expression may be confounded by factors such as age and sex.

In addition to these unique delirium-associated proteins, we identifid six upregulated proteins with a log2FC > 0.5 - PIGR, MST1, LBP, CRP, SAA1 and “SAA1;SAA2” - affecting different human body systems. After binding to the polymeric immunoglobulin receptor (PIGR), immunoglobulin (Ig) A and IgM are transported through epithelial cells to initiate an immune response(87, 88). This process has been studied for vaccination against the Middle East Respiratory Syndrome Coronavirus (89) and potential treatments against SARS-CoV-2 (90). However, it is speculative if this increased expression level could result from a more severe COVID-19 infection, as reflected by the higher SOFA score at admission in our patients with delirium.

The gene macrophage stimulating 1 (MST1) encodes a protein, also known as macrophage stimulating protein (MSP) or hepatocytes growth factor-like protein (HGFL), that interacts with the receptor tyrosine kinase recepteur d’origine nantais (RON) and activates its subsequent signal transduction pathway. This pathway has demonstrated the ability to maintain the blood-brain barrier in mice model(91). Additionally, Stephan and his colleagues suggested that this pathway interacts with the hippo pathway, which is associated with stress-related psychiatric disorders(92). Therefore, a potential association with delirium may be possible.

Inflammation is a major factor in the development of delirium(93). As such, C-reactive protein (CRP) is associated with delirium and can predict delirium, particularly in the perioperative setting (38, 94–96). Elevated CRP levels have been linked to poor outcomes and delirium(97). This could explain why we also discovered higher CRP levels in our patients with delirium.

Inflammation may also play a role in the protein function of serum amyloid A (SAA). This protein belongs to a group of polymorphic proteins, including SAA1 and SAA2. It is an acute-phase protein involved in lipid metabolism and is particularly associated with the surface of high-density lipoprotein (HDL). SAA has been linked to several diseases, such as sarcoidosis, atherosclerosis (98), and Alzheimer’s disease (99). Interestingly, Wiredu et al. showed that their postoperative patients with delirium had increased levels of SAA1 and SAA2(63), too. Our findings could support the hypothesis of a potential relationship between delirium and this specific protein family.

The colleagues Messner et al. have identified that the lipopolysaccharide-binding protein (LBP) is an associated factor for COVID-19 severity(12). It is also known that COVID-19 severity and delirium are connected, as reported by Kotfis(8). Considering this, we found that there is an increased level of LBP in patients with delirium. As LBP, an acute-phase protein, is involved in the defense against gram-negative bacteria(100), potential bacterial superinfections could also be an associated reason for delirium development in our patients.

Additionally, we found that only three proteins were downregulated with a log2FC < −0.5 in our study: pro-platelet basic protein (PPBP), haptoglobin (HP), and haptoglobin-related protein (HPR). PPBP, also known as platelet basic protein (PBP), is essential in defending the body against bacterial and fungal infections. Here, it acts as a chemoattractant and activator of neu-to SARS-CoV-2 by activating neutrophil recruitment through the network of pro-inflammatory C-X-C motif chemokine (CXCL) 6 and CXCL10-12 (102). A decreased level of PPBP may be a marker of COVID-19 severity, though its potential association with delirium remains unclear and is currently speculative.

### Pathway enrichment analysis of delirium-influenced proteins

In our study, we identified three significant pathways: the “Network map of SARS-CoV-2 signaling” (M42569/WP5115), “Acute inflammatory response” (M10617), and “Regulation of defense response” (M15277). These pathways involve inflammatory or host defense mechanisms and include overlapping proteins. Notably, two proteins were uniquely identified in the defense response pathway: SERPING1 and hemopexin (HPX). SERPING1 was found to be significantly upregulated in delirium patients, suggesting its potential involvement. There is limited evidence for hemopexin’s role in COVID-19 patients. De Lima et al. observed a non-significant elevation of hemopexin levels in COVID-19 patients compared to controls(103). The authors proposed that hemopexin might be a potential biomarker for COVID-19 severity. Our findings support this consideration, but further research is necessary to elucidate this potential connection.

*Proteome signature in comparison to other cohorts within the Pa-COVID 19 study*.

Delirium is positively associated with illness severity, as indicated by the SOFA score(104). At ICU admission, patients with delirium were more frequently observed to have higher SOFA scores than those without [non-delirium vs. delirium group; median (IQR); 5 (3–6) vs. 8 (7–12)]. Illness severity can also alter the proteome signature, as demonstrated by Messner et al. in a similar patient group within the Pa-COVID study(12). Interestingly, while many proteins were similarly up- or downregulated across both studies, we also identified distinct protein patterns unique to our delirium cohort. Specially, PIGR and MST1 appeared uniquely relevant in Messner et al.’s study. Conversely, HP was downregulated in our delirium group, but upregulated in cases of higher disease severity in the comparative study. Overall, severity of illness influences the proteome signature, yet delirium appears to shape its unique pattern within this profile.

### Strengths and limitations

The conduction of our study has several strengths. First, our study group investigated the proteome signature of COVID-19 patients over a more extended treatment period with multiple blood sampling time points. Second, the high-throughput mass spectrometry on undepleted human plasma and accompanied data analyzing platform(12) allowed us to explore a large scale of proteins simultaneously with similar proteomic depth as other contemporary mass spectrometry technologies. Third, the association of several new protein candidates with delirium presents an enhanced opportunity to better understand the pathomechanism of delirium in COVID-19 patients.

We need to acknowledge that our study has also several limitations. First, it was conducted in only one center and included only patients with COVID-19. As a result, inflammatory pathways might be enriched and could interact with other potential pathways. Therefore, our results cannot be generalized to other patient cohorts in the ICU. Second, since the proteome measurements were derived from blood samples, the findings may reflect systemic processes rather than directly representing brain metabolism or synaptic function. Third, in cases where the CAM-ICU was not performed, our approach to detecting delirium based on medical records could have resulted in inaccurate delirium status. Fourth, the limited number of quantified proteins available for enrichment analysis resulted in an incomplete interpretation and potential bias. Fifth, generalizing the functional context of proteins based on their UniProt gene names oversimplifies their biological process. Finally, although our included patients did not have neurological comorbidities, neurological complications could occur during the ICU treatment, leading to additional confounders of this study. Future work should also screen for neurological complications during the study to address this issue. These limitations underline the exploratory and hypothesis-generating nature of this study.

How applicable are our results to future research? This is an essential question because unbiased omics measurements are increasingly used to identify new biomarkers and pathomechanisms. However, our study cohort consists only of COVID-19 patients, an infectious disease that predominantly triggers inflammatory pathways. Moreover, specific COVID-19 treatments, including certain medication, may have influenced our results. Since delirium has more precipitating factors beyond infection, our findings are not generalizable to all potential pathogenesis of delirium.

## Conclusion

To the best of our knowledge, this study is the first to investigate proteomic signatures of COVID-19 patients with delirium in an ICU setting. Using high-throughput mass spectrometry, we demonstrated that a majority of proteins were associated with delirium and that a unique protein signature for delirium exists. These results may lead to further exploration of potential future clinical biomarkers to identify patients at risk of delirium, and uncover underlying etiologies as well as predisposing brain vulnerabilities. This can provide initial indications of treatments and their monitoring options. Our identification of both up- and downregulated proteins offers new insights into the pathomechanism of delirium. However, future studies involving a broader patient population and integrating additional omics approaches are essential to deepen our understanding of delirium pathophysiology.

## List of abbreviations

A2M: Alpha-2-Macroglobulin
“A2M; PZP”: Alpha-2-Macroglobulin and Pregnancy Zone Protein
AHSG: Alpha-2-HS-Glycoprotein (Fetuin-A)
APOM: Apolipoprotein M
APOM: Apolipoprotein M
ARDS: Acute Respiratory Distress Syndrome
BPS: Behavioral Pain Scale
BMI: Body Mass Index
C1INH: C1 Inhibitor
C7: Complement Component 7
CAM-ICU: Confusion Assessment Method for the ICU
CCA: Canonical Correlation Analysis
CD14: Cluster of Differentiation 14
CD5L: CD5 Antigen-Like Protein
CRRT: Continuous Renal Replacement Therapy
CRP: C-Reactive Protein
CST3: Cystatin C
COVID-19: Coronavirus Disease 2019
CPOT: Critical Care Pain Observation Tool
CXCL1: C-X-C Motif Chemokine
ECMO: Extracorporeal Membrane Oxygenation,
F2: Coagulation Factor II (Prothrombin)
F12: Coagulation Factor XII (Hageman Factor)
FETUB: Fetuin-B
FSAP: Factor VII Activating Protease
GCS: Glasgow Coma Scale
HA: Hyaluronic Acid
HABP2: Hyaluronan-Binding Protein 2
HDL: High-Density Lipoprotein
HGFL: Hepatocytes Growth Factor-Like Protein
HMWK: High Molecular Weight Kininogen
HP: Haptoglobin
“HP; HPR”: Haptoglobin and Haptoglobin-Related Protein
HPX: Hemopexin
ICU: Intensive Care Unit
Ig: Immunoglobulin
IGFALS: Insulin-Like Growth Factor-Binding Protein, Acid Labile Subunit
IGHA1: Immunoglobulin Heavy Constant Alpha 1
IGHV: Immunoglobulin Heavy Chain Variable region
ITIH3: Inter-Alpha-Trypsin Inhibitor Heavy Chain 3
ITIH4: Inter-Alpha-Trypsin Inhibitor Heavy Chain 4
IQR: Interquartile Range
KKS: Kallikrein-Kinin-System
KLKB1: Kallikrein B, Plasma (Fletcher Factor) 1
KNG1: Kininogen-1
LBP: Lipopolysaccharide-Binding Protein
LMWK: Low Molecular Weight Kininogen
Log2FC: Log2 Fold-Change
MSP: Macrophage Stimulating Protein
MST1: Macrophage Stimulating 1
NTI: Non-Thyroidal Illness
NRS: Numerical Rating Scale
PBP: Platelet Basic Protein
PIGR: Polymeric Immunoglobulin Receptor
PPBP: Pro-Platelet Basic Protein
PRG4: Proteoglycan 4
RASS: Richmond Agitation Sedation Scale
RON: Receptor Tyrosine Kinase Recepteur D’origine Nantais
RT-PCR: Reverse Transcriptase Polymerase Chain Reaction
SAA (1): Serum Amyloid A (1)
“SAA1; SAA2”: Serum Amyloid A1 and Serum Amyloid A2
SARS-CoV-2: Severe Acute Respiratory Syndrome Coronavirus 2
SERPINA7: Serpin Family A Member 7 (Thyroxin-Binding Globulin)
SERPING1: Serpin Family G Member 1 (C1 Inhibitor)
SOFA: Sequential Organ Failure Assessment
SOP: Standard Operating Procedure
TBG: Thyroxin-Binding Globulin
TGFBI: Transforming Growth Factor-Beta-Induced Protein
TGF-β: Transforming Growth Factor – β
T3: Triiodothyronine
T4: Thyroxine
TSH: Thyroid-Stimulating Hormone
WHO: World Health Organization

## Declarations

### Author contributions

FK, MR, and CS designed the study. The Pa-COVID-19 study group including AE did data and sample collection. MR, VD, MM performed protein profiling. JS and AE analyzed the data. FK, MR, EB, JS, AE and CS interpreted the data. AE and JS wrote the first draft of the paper. All authors reviewed and contributed to finalizing the manuscript.

## Data Availability

The datasets analyzed during the current satellite study are available from the corresponding author on reasonable request. The table of primary protein quantities together with the corresponding metadata, has been formerly published at https://doi.org/10.1371/journal.pdig.0000007. Additionally, the mass spectrometry proteomics data have been submitted to the ProteomeXchange Consortium (http://proteomecentral.proteomexchange.org) via the PRIDE partner repository and assigned the dataset identifier PXD029009.

https://proteomecentral.proteomexchange.org/cgi/GetDataset?ID=PXD029009

## Acknowledgements

PA-COVID-19 Study group, Charité–Universitätsmedizin Berlin

Malte Kleinschmidt, Katrin M. Heim, Belén Millet, Lil Meyer-Arndt, Nils B. Müller, Ralf H. Hübner, Tim Andermann, Jan M. Doehn, Bastian Opitz, Birgit Sawitzki, Daniel Grund, Peter Radünzel, Mariana Schürmann, Thomas Zoller, Fridolin Steinbeis, Florian Alius, Philipp Knape, Astrid Breitbart, Yaosi Li, Felix Bremer, Panagiotis Pergantis, Susanne Fieberg, Anne Wetzel, Moritz Müller-Plathe, Timur Özkan, Carola Misgeld, Dirk Schürmann, Bettina Temmesfeld-Wollbrück, Britta Stier, Martin Möckel, Jan A. Graaw, Victor Wegener, Marc Kastrup, Felix Balzer, Daniel Wendisch, Sophia Brumhard, Sascha S. Haenel, Philipp Georg, Claudia Conrad, Kai-Uwe Eckardt, Lukas Lehner, Jan M. Kruse, Carolin Ferse, Roland Körner, Steffen Weber-Carstens, Alexander Krannich, Saskia Zvorc, Linna Li, Uwe Behrens, Sein Schmidt, Maria Rönnefarth, Christina Pley, Claudia Fink, Chantip Dang-Heine, Robert Röhle, Emma Lieker, Christian Wollboldt, Yinan Wu, Georg Schwanitz, Constanze Lüttke, Denise Treue, Michael Hummel, Victor M. Corman, Christian Drosten, Christof von Kalle

## Notes

### Competing Interest Statement

Andreas Edel declares that his institution received payments or honoraria for lectures and expert testimony from Gilead Sciences GmbH and Draeger Medical Deutschland GmbH. Andreas Edel hold shares of BioNTech and Novavax (less than 5000 Euro). Andreas Edel declares to be a member of the German Society of Anaesthesiology and Intensive Care Medicine (DGAI) and participate in the DGAI Round Table on Tele ICU. Additionally, he collaborates in the section on Metabolism and Nutrition of the German Interdisciplinary Association for Intensive and Emergency Medicine (DIVI). He holds memberships in several other organizations, including the German Sepsis Society (DSG), the European Society of Intensive Care Medicine (ESICM), the European Society of Anaesthesiology and Intensive Care (ESAIC), the German Society for Nutritional Medicine (DGEM), and the European Society for Clinical Nutrition and Metabolism (ESPEN), although he is not actively involved in committee work in these organizations. Jayanth Sreekanth declares that he has no competing interests. Florian Kurth received a research grant from GSK Inc. for unrelated work. He has also received honoraria for lectures from Pfizer Inc. and was supported by Gilead Inc. to attend a conference. Markus Ralser is founder, CSO and shareholder of Elitptica Ltd. There is no relationship of this role and the current work. Vadim Demichev reports receiving funding from the German Federal Ministry of Education and Research (BMBF) under research grant 161L0221 in support of this manuscript and declares his shareholder interest in Aptila Biotech GmbH. Michael Muelleder reports receiving funding from the German Federal Ministry of Education and Research (BMBF) under research grant 16LW0239K in support of this manuscript. He is a consultant and shareholder at Eliptica Ltd. He also received financial support from Thermo Scientific to attend the HUPO 2024 conference, which covered conference fees for a presentation given at their on site User Meeting. Additionally, he holds patents planned, issued, or pending in association with Inoviv, Inc., including a joint patent for a COVID 19 Multiple Reaction Monitoring (MRM) panel. Eric Blanc declares that he has no competing interests. Claudia Spies reports grants from The European Commission Horizont Europa, German Federal Joint Committee (Gemeinsamer Bundesausschuss GBA), German Federal Ministry of Education and Research (BMBF), Philips Electronics Nederland BV, Max Planck Society, Sintetica GmbH, Dr. F. Koehler Chemie GmbH, Georg Thieme Verlag, German Federal Ministry for Economic Affairs and Climate Action (BMWI), European Society of Anesthesiology and Intensive Care, Stifterverband (non profit society promoting science and education), Charite internal university grants, Einstein Foundation Berlin, German Aerospace Center (Deutsches Zentrum fuer Luft und Raumfahrt e.V. DLR) and German Research Society (Deutsche Forschungsgemeinschaft) during the conduct of the study. In addition, Claudia Spies has different patents and an unpaid leadership or fiduciary role in association of the Scientific Medical Societies in Germany (AMWF), German Research Foundation (Deutsche Forschungsgemeinschaft), German National Academy of Sciences Leopoldina (Deutsche Akademie der Naturforscher Leopoldina e. V.), Berlin Medical Society (Berliner Medizinische Gesellschaft), ESICM European Society of Intensive Care Medicine, ESAIC European Society of Anaesthesiology and Intensive Care, German Society of Anaesthesiology and Intensive Care Medicine (Deutsche Gesellschaft fuer Anaesthesiologie und Intensivmedizin DGAI), German Interdisciplinary Association for Intensive Care and Emergency Medicine(Deutsche Interdisziplinaere Vereinigung fuer Intensiv und Notfallmedizin DIVI) and German Sepsis Foundation (Deutsche Sepsis Stiftung). Claudia Spies has participated on Data Safety Board or Advisory boards for Prothor, Takeda Pharmaceutical Company Limited and Lynx Health Science GmbH.

### Clinical Protocols

https://pmc.ncbi.nlm.nih.gov/articles/PMC7293426/

### Funding Statement

The study was further supported by the German Federal Ministry of Education and Research (NaFoUniMedCovid19 NUM NAPKON, NUM COVIM, FKZ: 01KX2021 and PROVID FKZ: 01KI20160A) as well as BIH and Charite internal research funds.

### Author Declarations

The study was approved by the ethics committee of the Charite-Universitaetsmedizin Berlin under the reference number EA2/066/20.

### Summary of Updates

The model was previously mischaracterized; this has now been corrected.

