## supplementary figures and tables for "Unique proteome signatures in ICU patients with COVID-19 and delirium: an observational study": Supplementary.docx

**Supplementary Figure 1** **Heatmap of patient stays in the hospital.** A graphical representation of delirium occurrence during the hospital stays of each patient in the study. Day 0 in the x-axis corresponds to patients’ admission day, and the color-coded blocks represent ICU or hospital treatment. The days when blood samples were drawn for protein analysis are marked using a black dot inside the color-coded blocks. The blank cells indicate that the patient was either treated in the normal ward or not in the hospital. Red- and sea-green-colored blocks refer to Delirium and Delirium-free days respectively.

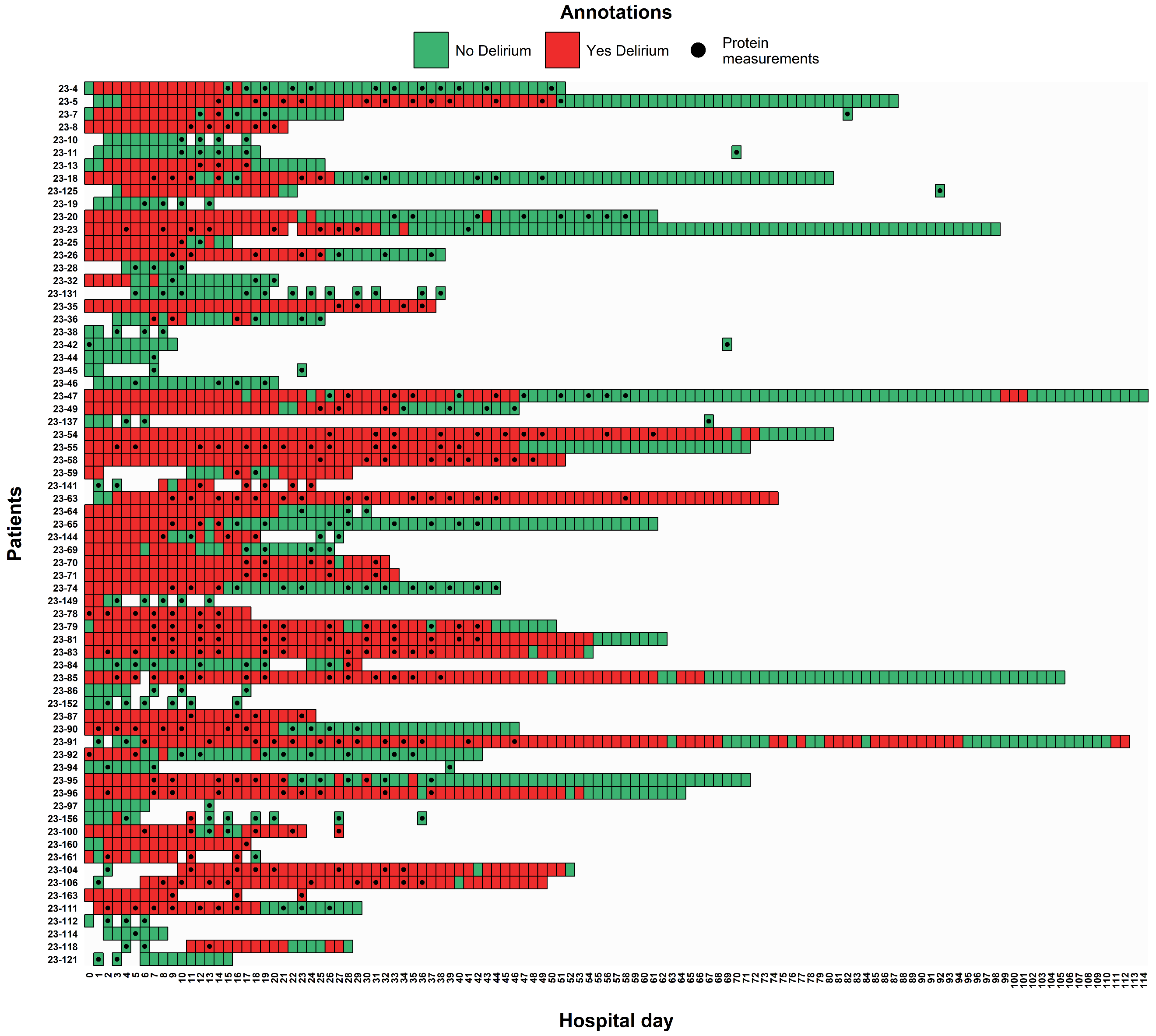

**Supplementary File 1:** Venn_Diagram_tables.xls

The supporting file to the Venn Diagram in Figure 2, containing unique and common significant proteins between the model coefficients.

**Supplementary File 2:** Differential_expression_results.xls

Differential expression of proteins from the multivariate regression model when delirium, age, and sex were defined as coefficients and when the variables were defined as model coefficients separately.

**Supplementary File 3:** Enrichment_results.xls

Table of pathways enriched for proteins differentially expressed according to delirium.
